# Modelling-based evaluation of the effect of quarantine control by the Chinese government in the coronavirus disease 2019 outbreak

**DOI:** 10.1101/2020.03.03.20030445

**Authors:** Xinkai Zhou, Zhigui Wu, Ranran Yu, Shanni Cao, Wen Fang, Zhen Jiang, Fang Yuan, Chao Yan, Dijun Chen

**Affiliations:** State Key Laboratory of Pharmaceutical Biotechnology, School of Life Sciences, Nanjing University, Nanjing 210023, China; Institute of Artificial Intelligence Biomedicine, Nanjing University, Nanjing 210023, China

**Author notes:** These authors contributed equally to this work (Xinkai Zhou, Zhigui Wu, Ranran Yu, Shanni Cao).

**Keywords:** COVID-19, retrospective modeling, simulation, epidemic spreading, transmission model

## Abstract

The novel coronavirus disease 2019 (COVID-19) epidemic, which was first identified in Wuhan, China in December 2019, has rapidly spread all over China and across the world. By the end of February 2020, the epidemic outside Hubei province in China has been well controlled, yet the next wave of transmission in other countries may have just begun. A retrospective modeling of the transmission dynamics would provide insights into the epidemiological characteristics of the disease and evaluation of the effectiveness of the strict measures that have been taken by central and local governments of China. Using a refined susceptible-exposed-infectious-removed (SEIR) transmission model and a new strategy of model fitting, we were able to estimate model parameters in a dynamic manner. The resulting parameter estimation can well reflect the prevention policy scenarios. Our simulation results with different degrees of government control suggest that the strictly enforced quarantine and travel ban have significantly decreased the otherwise uncontrollable spread of the disease. Our results suggest similar measures should be considered by other countries that are of high risk of COVID-19 outbreak.

**Summary:** *Background:* The novel coronavirus disease 2019 (COVID-19) epidemic, which was first reported in Wuhan and rapidly spread across the world, has been well controlled in China but is only starting to take off in other countries. Here we provide a retrospective modelling analysis of the transmission dynamics in China and evaluated the effectiveness of the strict government control strategies.

*Methods:* We considerably refined the original susceptible-exposed-infectious-removed (SEIR) transmission model, and used the publicly available data from Jan 13^rd^ to Feb 29^th^ for model fitting and parameter estimation in a dynamic manner considering effect of prevention policies. We then used the estimated model parameters to simulate the epidemic trend and transmission risk of the disease with various degrees of government control.

*Findings:* The severity rate and the fatality rate remain unchanged during the whole epidemic. While government intervention had a moderate effect on the incubation rate (σ), the recovered rate (γ) endured several fold increase. Strikingly, a significant decrease in the infectious rate (β) was observed. Without government control, peak infected cases in Wuhan would reach 7.78 million (70% of the whole population) and total deaths could reach 319000 based on the current mortality rate (4.1%).

*Interpretation:* Our simulation results with different degrees of government control suggest that the strictly enforced quarantine and travel ban have significantly decreased the otherwise uncontrollable spread of the disease. Our results suggest similar measures should be considered by other countries that are of high risk of COVID-19 outbreak.

*Funding:* The National Natural Science Foundation of China (21877060).

*Research in context:* Evidence before the study
A global outbreak of coronavirus disease 2019 (COVID-19), caused by the severe acute respiratory syndrome coronavirus 2 (SARS-CoV-2), has been posing significant threats to public health worldwide. By the end of February 2020, 87645 confirmed cases are reported around the world, including 7330 severe cases and 2994 fatalities. We searched PubMed and preprint archive for papers published up to Feb 29^th^, 2020, using keywords “COVID-19”, “SARS-CoV-2”, “2019-nCoV”, and “novel coronavirus.” We found several researches on the transmission dynamics of COVID-19; however, only one preprint predicted the effect of government intervention in China with incomplete epidemiological data. Added value of this study
Since the epidemic is already close to its end in China except Wuhan city, we have the opportunity to carry out a relatively complete retrospective analysis. We optimized the SEIR model using a dynamic fitting approach, taking into account the government measures and reached a much more precise fitting of the data comparing to other studies published. We showed that the severity rate and the fatality rate remain unchanged during the whole epidemic, suggesting the only effective way to control the disease is to control the number of infections. While government intervention had a moderate effect on the incubation rate (σ), it is essential for increasing the recovered rate (γ), and for decreasing and stabilizing the infectious rate (β). We also simulated the scenarios with various degrees of government control which could be a useful tool to predict the necessity of government intervention. An interactive online application was made available to the public on Feb 24^th^, 2020. Implications of all the available evidence
The COVID-19 outbreak has already been effectively controlled in China; however, the risk of rapid global explosion is extremely high due to the high transmissive rate of the SARS-CoV-2 virus. The quarantine measures adopted by the Chinese government are essential for the control of the COVID-19 epidemic.

## Introduction

The recent outbreak of novel coronavirus disease 2019 (COVID-19) has already become an epidemic of global scale. Two months ago, on Dec 30^th^ 2019, the first confirmed case was officially reported by local authorities of Wuhan city, China^1,2^. The pathogen was soon identified as a new coronavirus on Jan 7^th^ 2020 and tentatively named 2019-nCOV^2,3^ and later renamed SARS-CoV-2 by the International Committee on Taxonomy of Viruses^4^. However, strict control measures, including the lockdown of Wuhan city did not take effect until Jan 23^rd^, by which time several million people have already travelled outside Wuhan for the upcoming Chinese New Year, spreading the disease all over China. By the end of January, confirmed cases were reported by several other countries, leading to the declaration of Public Health Emergency of International Concern (PHEIC) by WHO^5^ on Jan 30^th^. For the whole month of February, rigorous prevention and control measures were taken by the Chinese government and the whole country is literally in a lockdown state. The necessity of these strict measures has been questioned both domestically and internationally concerning the devastating effects on the global economy; and there is also severe debate over when these control measures, including travel bans and public gathering bans, should be lifted. However, the effectiveness of these strict control measures has not been systematically evaluated using epidemiological data.

Here we provided a retrospective modelling-based evaluation of the effectiveness of government control measures on COVID-19 in China. Using a well-recognized susceptible-exposed-infectious-recovered (SEIR) model, we fitted the public data by taking the effect of prevention policies into consideration and achieved an almost perfect fit for the real data. Key epidemiological parameters including incubation rate (σ), infectious rate (β) and recovered rate (γ) and were estimated in a dynamic manner. These estimates were then used to parameterize our model and different outcome scenarios were simulated with various degrees of government control. An interactive web-based Shiny application based on our modelling is now available online (http://compbio.nju.edu.cn/ncov2019/). Our findings may provide a useful tool for analyzing the impact of control measures and predicting the transmission dynamics in other countries.

## Methods

### Data collection

Daily cumulative number of confirmed cases, dead cases and recovered cases (from Jan 13^rd^, 2020 to Feb 29^th^, 2020) infected by COVID-19 were collected from the Tencent social networks (https://news.qq.com/zt2020/page/feiyan.htm); the daily number of severe cases were obtained from official websites of the National Health Commission of China and of health commissions of provinces, municipalities and major cities. Overseas data were obtained from the official website of WHO (https://www.who.int/emergencies/diseases/novel-coronavirus-2019/situation-reports/). All collected data were double checked. By finalizing this manuscript, the period of data coverage starts from mid-January to the end of February 2020.

### Model, parameter estimation and simulation

The susceptible-exposed-infectious-recovered (SEIR) compartmental model is one of the best models to describe the epidemic of diseases with a latent phase like the SARS-CoV-2. We adopted the following ordinary differential equation (ODE) model to simulate the epidemic of COVID-19 for each investigated population:

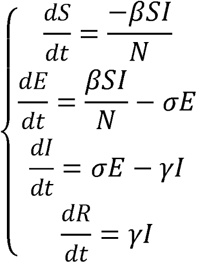

where S, E, I, and R were the number of susceptible, latent (or exposed), infectious, and removed individuals (including recovered individuals and death) at time t and N − S + E + I + R is the size of population. We assume that N is constant in a investigated population. In the SEIR model, the infectious rate, β, controls the rate of spread which represents the probability of transmitting disease between a susceptible and an infectious individual. The incubation rate, σ, is the rate of latent individuals becoming infectious (average duration of incubation is 1/σ). Recovery rate, γ, is the rate of infectious individuals removed from the transmission system, which is determined by the average duration of infection.

We suppose that the epidemic spreading trend of COVID-19 suffered the first period of free propagation (before Jan 23^rd^, 2020) and then under the health policies intervention. Therefore, there is no one-fit-all set of parameters to fit the model with the aggregate data. Instead, we assume that the model parameters dynamically changed during the epidemic course and was dependent on the degree of prevention. Based on such an assumption, we inferred the model parameters in a stepwise manner at each time point, using the latest officially confirmed infected data. We then investigated the pattern of parameter dynamics and correlated these with various prevention measures in different populations. As expected, the pattern of estimated parameter (especially the infectious rate β) dynamics can well reflect the outcome of control policies. We further forecasted the epidemic spreading by estimating the model parameters using polynomial regression-based machine-learning approach, which has been used to describe nonlinear phenomena such as the progression of disease epidemics^6^. Finally, we simulated possible spreading course of COVID-19 in Wuhan subjected to different quarantine rate as reflected by the dynamics of the predicted parameters.

### Statistical analysis

If not specified, all statistical analyses and data visualization were done in R (version 3.6.2). We used non-parametric tests to assess differences among different group (Wilcoxon test to compare two groups and Kruskal-Wallis test to compare three or more groups). We used R packages such as ggplot2 and plotly for graphics.

## Results

Since the impact of an epidemic depends on both the number of persons infected and the spectrum of clinical severity, we first analyzed the change of severity rate and the fatality rate over time and found that both parameters remain unchanged over the investigated phase for each province and are not affected by the strictness of control measures (**Fig. 1**), indicating that total number of severe cases and deaths are largely determined by the total number of infections, thus supporting the importance and urgency of outbreak prevention and control of transmission.

**Figure 1.**
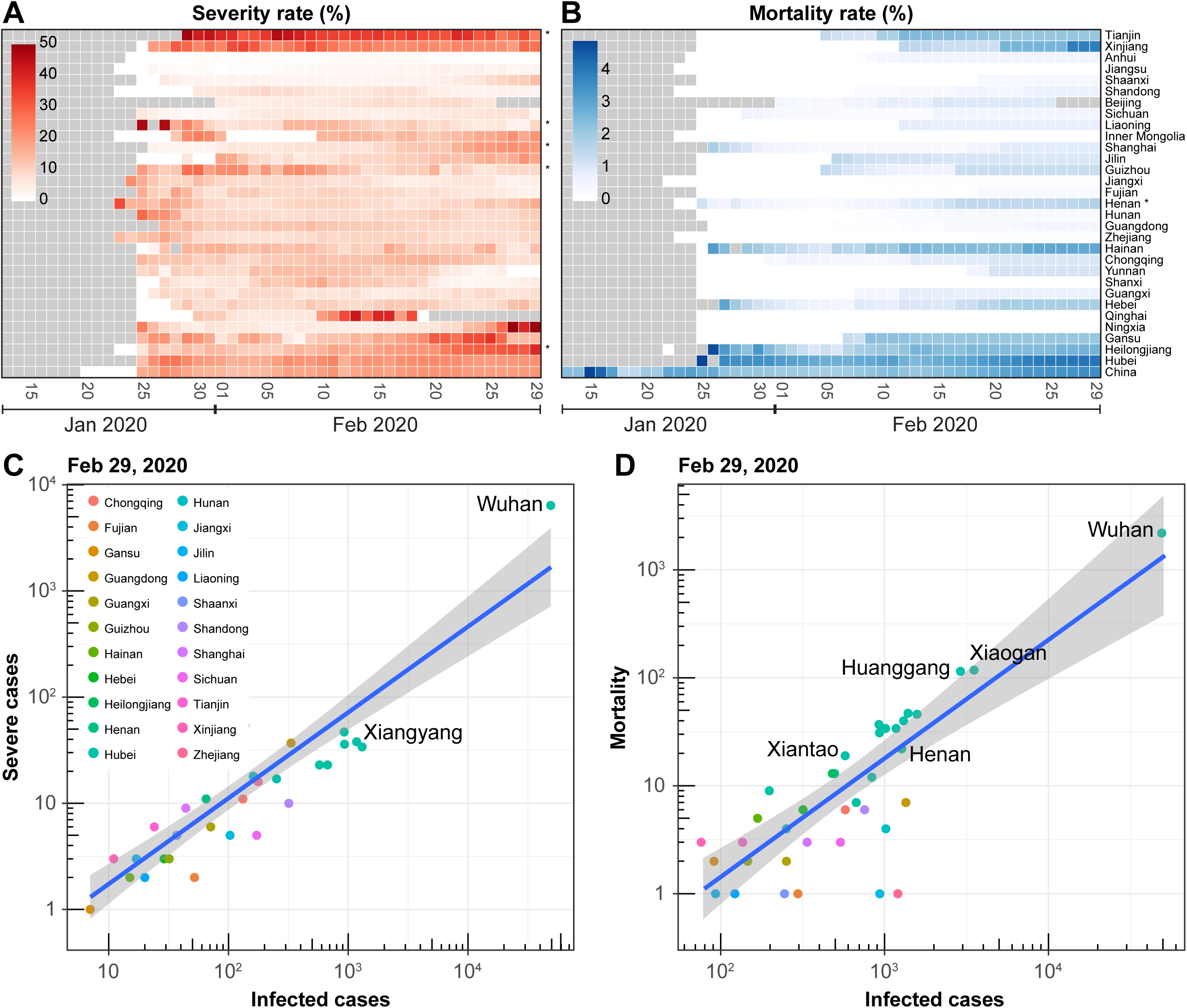
The severity rate and themortality rate of COVID-19 in mainland China from Jan 13^rd^ to Feb 29^th^, 2020. Heatmaps showing the severity ratio **(A)** and the mortality rate **(B)** in different provinces or municipalities of mainland China. The monotonic distribution of data over time was assessed by Mann-Kendall trend tests. P-values adjusted were corrected with the Benjamini-Hochberg method in order to control the false discovery rate. “*” indicates an adjusted p-value < 0.001. Scatter plots showing the positive correlation of the number of severe cases **(C)** or mortality **(D)** and infected cases on Feb 29^th^, 2020.

The susceptible-exposed-infectious-recovered (SEIR) transmission model is one of the best models to describe the epidemic of diseases with a latent phase. However, it is only accurate under conditions of no intervention. In the real world, various degrees of intervention have been adopted; as a result, the SEIR model does not truly reflect the spread of the epidemic. To solve this discrepancy, we optimized the model in a stepwise manner at each time point, using the latest official confirmed infected data to fit the SEIR model. We thus obtained dynamic changes of the three model parameters: infectious rate (β), incubation rate (σ) and recovered rate (γ) (**Fig. 2A**). As expected, incubation rate (σ) shows little to none change over time (**Fig. 2B**,**C**). In this regard, the average duration of incubation is estimated to be 5.9 days based on data in Wuhan, which is generally in agreement with existing reports^7–10^. While recovered rate (γ) shows a slow but steady increase over time (**Fig. 2D**), the infectious rate dramatically changed during the whole outbreak process (**Fig. 2B**,**E**). The national infectious rate clearly peaked around Jan 23^rd^, the day of Wuhan lockdown and has continued to drop ever since. This trend is more apparent when analyzing the data of Wuhan city alone (**Fig. 2E**). The curve for other cities displayed a continuous descending trend from the beginning of the available data point (around Jan 20^th^, the city of Nanjing was shown as an example in **Fig. 2E**), when strict control measures have already been in place. In Wuhan, the recovery rate (γ) has continued to increase with more than 2-fold change which is probably due to the dramatic improvement of medical conditions, such as the immobilization of thousands of physicians from all over the country and immediate establishment of several new hospitals in Wuhan. These data indicate a significant correlation between the infectious rate (β) and the recovery rate (γ) with prevention and control measures.

**Figure 2.**
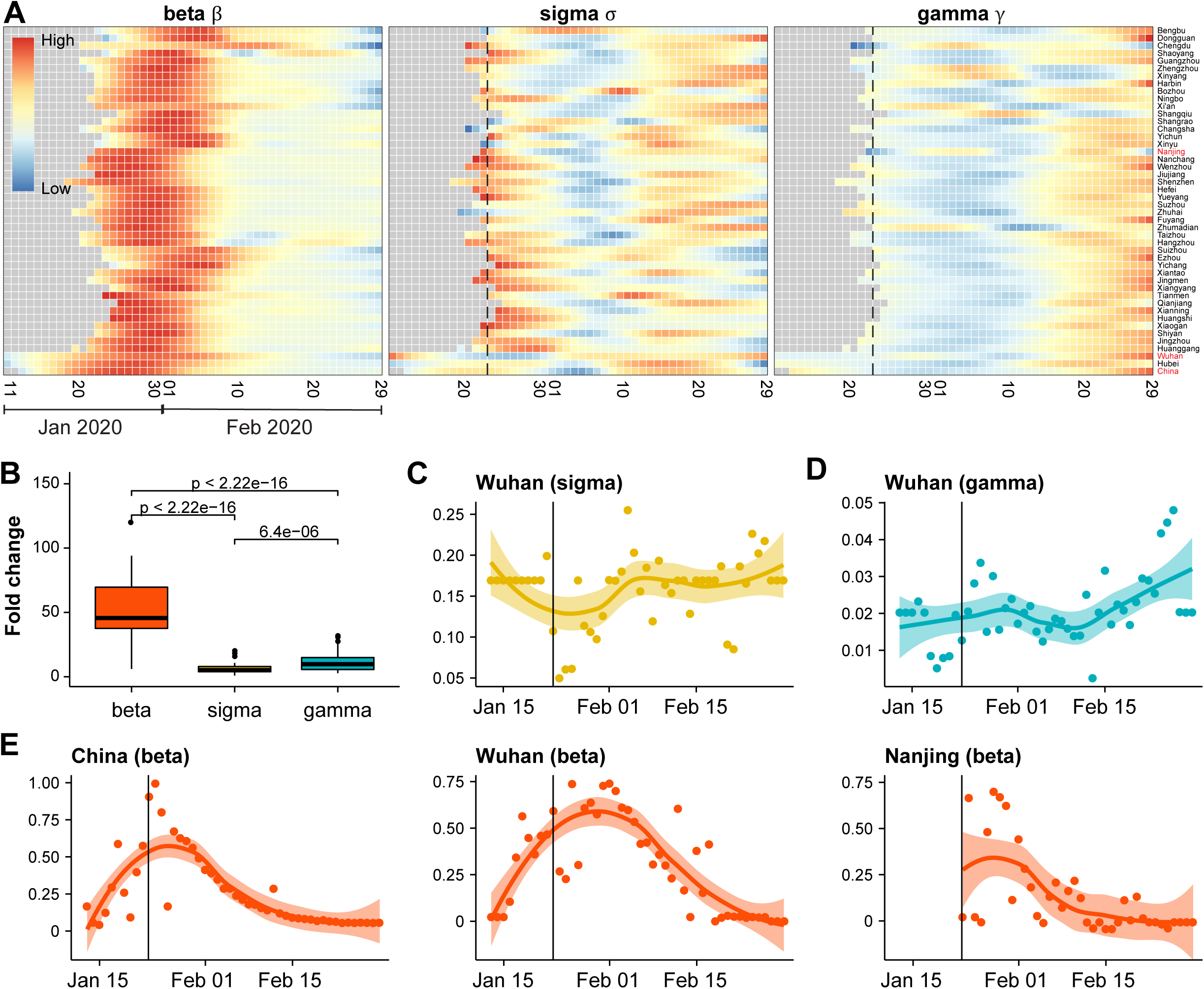
Parameter estimation of the SEIR transmission model. (A) Heatmap showing the dynamic change of estimated parameters of the SEIR model over time. In order to compare data among different cities, only relative values are shown in the heatmap. Data for example cities (in red) are shown in **(C-E). (B)** Fold changes of the three parameters of the SEIR model. Dynamics of estimated σ **(C)** or γ **(D)** parameters over time in Wuhan. **(E)** Dynamics of estimated β parameter over time in the population of mainland China (left), Wuhan city (middle) and Nanjing city (right). In this figure, the modeling analysis was performed in the populations of the Hubei province, mainland China as well as the top 44 cities of the whole mainland China. Dash lines indicate the date of lockdown of Wuhan.

Next, we simulated the possible outcome scenarios with different degrees of government prevention. Assuming there is a direct correlation with the value of infectious rate and the strictness of prevention measures (as shown in **Fig. 2**), we simulated the outbreak dynamics in different prevention scenarios using different estimated parameters in Wuhan (**Fig. 3A**). In the case of low to no prevention (with high infectious rate at the early stage), the peak of predicted infection cases would eventually reach ca. 7.78 million, covering 70% of the whole population of Wuhan city. The number of potentially infected individuals dramatically reduce as the infectious rate decreases upon prevention interventions (**Fig. 3B**). These data suggest that strict prevention is vitally important to reduce the peak of infection cases.

**Figure 3.**
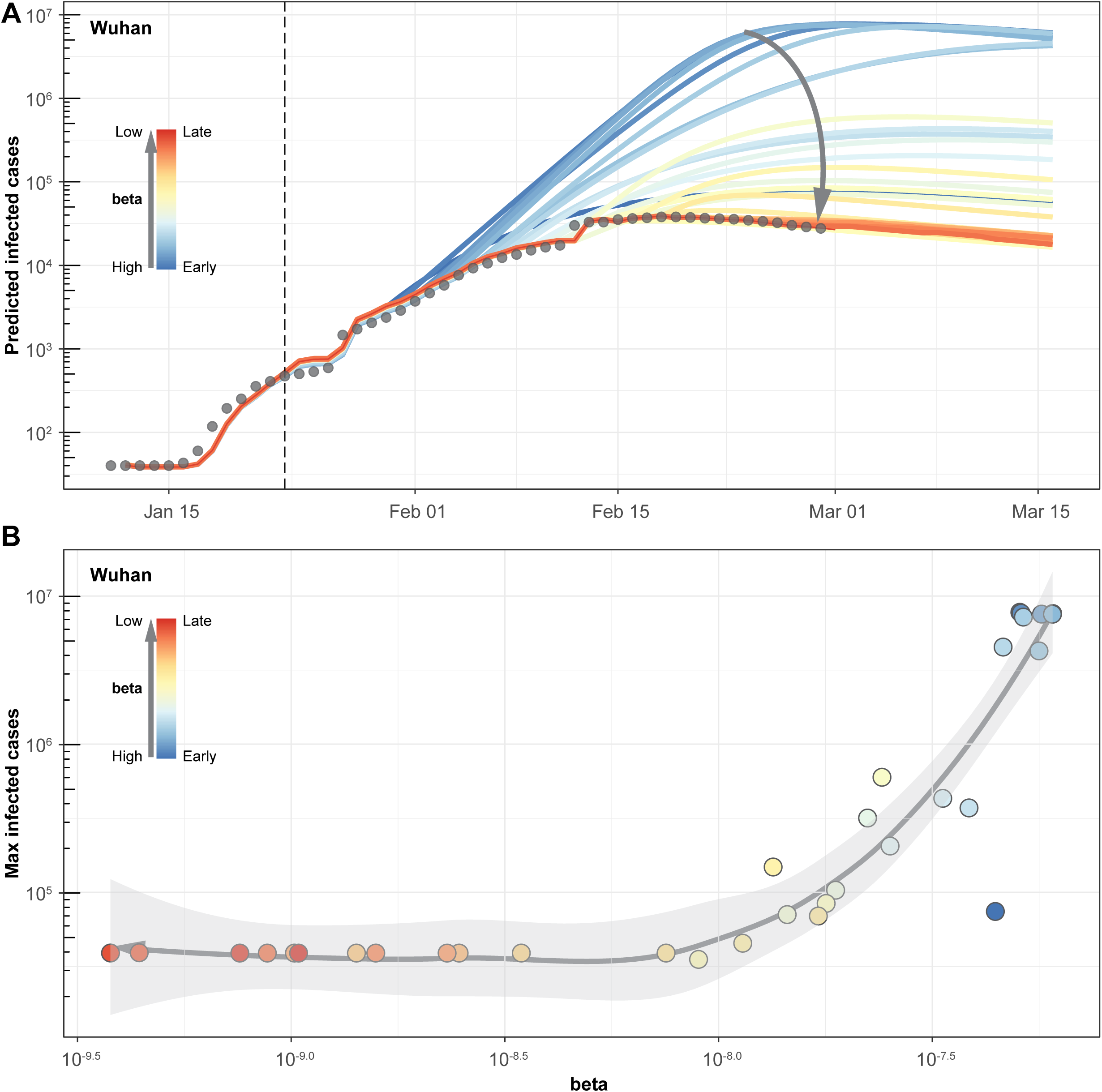
Simulation of epidemic spreading in Wuhan starting with different prevention scenarios. **(A)** Simulation analysis showing the predicted confirmed individuals based on estimated values of the β parameter at different time points. β values are high at the early stage and low at the late stage, as indicated in the color legend. Arrow shows the change of epidemic curve under different prevention scenarios. **(B)** Relationship between the estimated β parameter and the peak of predicted confirmed individuals using the corresponding β value.

Using a similar approach, we next modeled all the publicly available data on several different countries that already have considerable amounts of confirmed cases and predict their epidemic curve with various degrees of government control, assuming similar transmission properties of the virus in different counties (**Fig. 4**).In Singapore, where only mild government intervention existed, a surprisingly continuous decrease in the infectious rate (β) and increase in the recovery rate (γ) was observed, implying the potential inhibitory effect of warm climate on the spreading of the virus; whereas the situation in Italy, Korea, and Iran are quickly deteriorating. For Japan, although the infectious rate (β) is getting better, the recovery rate (γ) is not, implying either a latency in hospitalization or lack of dedicated medical resources.

**Figure 4.**
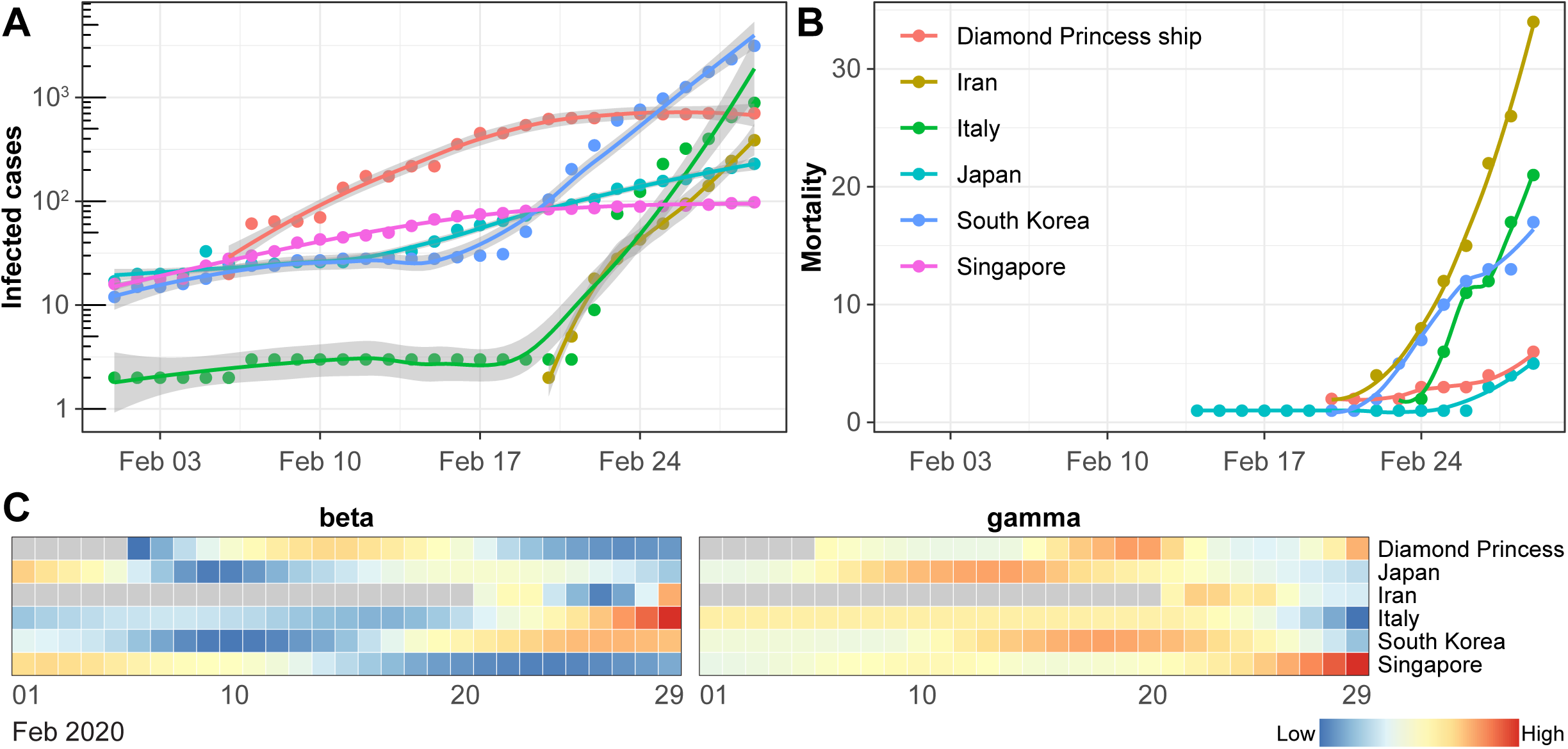
Analysis of the COVID-19 data outside of China, with a focus on the outbreak in February. The reported infected cases **(A)** and mortality **(B)** in the top five countries and the Diamond Princess ship in February 2020. Heatmaps show the dynamic change of estimated parameters β **(C)** and γ **(D)** of the SEIR model over time.

## Discussion

By the end of February 2020, two months after its outbreak in Wuhan, the COVID-19 epidemic has already spread to more than fifty countries. Areas with a high risk of exponential outbreak include Japan, South Korea, Italy, Singapore and Iran. Since the situation in China has already been well controlled, a retrospective evaluation of the epidemiological characteristics and transmission dynamics would provide valuable insights that might help with disease control decisions world-wide. To our knowledge, our work is the first to evaluate the effectiveness of government control on the spread of COVID-19 using a modelling approach.

In agreement with others^11–14^, we found the SEIR-like models to be the most appropriate model for analyzing the transmission dynamics of COVID-19. However, the traditional approach for fitting the SEIR model is based on the overall trend data of the epidemic using hypothesized parameters and thus the model do not well fit the real-life data (i.e. the real-life data trend does not conform to the ideal SEIR model curve). The main reason is that the traditional approach does not take into account the impact of prevention and control strategies on the epidemic dynamics. In this study, we adjusted the SEIR model to objectively reflect the impact of prevention and control strategy on the infectious rate. Nonetheless, our study has several major limitations. Firstly, we were not able to derive a generalized model with finalized parameters to fit the overall epidemic spreading. Instead, the model parameters were estimated in a dynamic manner in our study. This is in line with the observation of dynamic transmission properties of the virus^15^. Secondly, the significance of the model largely depends on the accuracy of estimated parameters. Although there is not a golden standard so far to evaluate the accuracy of our model parameters, the estimated duration of incubation (1/σ) from our model-derived parameter incubation rate (σ) is highly in agreement with previous estimations by other studies^7–10^, and the dynamic values of the infectious rate (β) is within the range of the recent estimation by Yang et al.^14^. These observations suggest our approach for model parameter estimation is plausible. Thirdly, our epidemic forecast was somehow sensitive to our prediction of parameters which were estimated from their overall dynamic patterns. Nonetheless, such forecasting would still be accurate in a short period. In this regard, artificial intelligence (AI)-inspired methods^14,16^ may be alternatives to epidemiological models for the real-time forecasting of transmission dynamics of COVID-19.

The fact that the severity and the fatality rate remain unchanged during the whole epidemic course suggests that the biology of the virus itself did not change over time; this is in line with genetic sequencing results that few mutations are identified among virus samples collected from different generations of patients^17,18^. However, it is hard to predict whether this feature of the virus will change as it spread in other countries. The significantly higher reported fatality rate in Wuhan city and Hubei province comparing to other regions are probably due to the shortage of medical supplies and the underestimation of total cases of infection. The lessons in Wuhan also taught us that if the outbreak in a heavily populated metropolitan area is not well controlled soon enough, rapid saturation of the hospital capacity is inevitable and devastating. Therefore, the only effective way is to control the epidemic is to mitigate transmission (infectious rate) and this has proved to be successful in China.

There is no doubt that the capability of management and control of COVID-19 transmission heavily rely on the preparedness of a country’s health system. While it remains debatable whether large economy bodies in Asia such as South Korea and Japan should adopt similar control measures as the Chinese government; in less developed countries with insufficient medical resources and absence of a pandemic preparedness plan, a mild response might be inadequate to deal with such an outbreak. Extreme quarantine and transport control measures similar to China should be considered to mitigate local transmission following confirmed importation.

In conclusion, we have constructed an SEIR model and fitted all the publicly available China COVID-2019 data in a dynamic manner, and get a fairly accurate model of the whole process of the epidemic in China. We estimated the overall fatality rate to be 0.68% outside of Hubei province. Through simulation, we also evaluated the importance and effectiveness of strict government control enforced by Chinese authorities. It is clear that the control measures, both in Wuhan and nationwide, have significantly reduced transmissibility. Considering the potential threat of fast worldwide COVID-19 outbreak to public health and global economy, more strict government controls are advised based on the China experience.

## Contributors

DC and CY designed the study. CY and DC did the literature search. XZ, ZW, RY and SC collected and manually checked the data. DC analyzed the data and developed the Shiny application with support from XZ, ZW, ZJ and WF. DC designed the figures. CY and DC interpreted the results and wrote the manuscript with input from FY.

## Data Availability

The original data that support the findings of this study are available from the National Health Commission of the People’s Republic of China as well as the WHO.

## Data sharing

The original data that support the findings of this study are available from the National Health Commission of the People’s Republic of China as well as the WHO. In order to efficiently use our compiled data as well as modelling results by external users, we developed an integrative web-based application with the Shiny framework (http://shiny.rstudio.com/), which combines the computational power of R with friendly and interactive web interfaces. Our compiled database is publicly available from the website of Nanjing University (http://compbio.nju.edu.cn/ncov2019/) on Feb 24^th^, 2020, where it would be updated in time during the outbreak. Data used in this analysis is frozen on Feb 29^th^, 2020.

## Acknowledgments

We would like to thank Chenyu Zhang from Nanjing University (NJU) and Yu Xue from Huazhong University of Science and Technology (HUST) for helpful discussion and suggestions. We thank Wanshan Ning from HUST, and Yikai Zhu and Liang-Yu Fu from NJU for technical support. The study was supported by the Fundamental Research Funds for the Central Universities and the National Natural Science Foundation of China (21877060).

